# Re-evaluating malarial retinopathy to improve its diagnostic accuracy in cerebral malaria

**DOI:** 10.1101/2024.12.17.24319176

**Authors:** Kyle J Wilson, Alice Muiruri Liomba, Karl B Seydel, Christopher A Moxon, Ian JC MacCormick, Simon P Harding, Nicholas AV Beare, Terrie E Taylor

## Abstract

**Background:** Previous work has identified that malarial retinopathy (MR) has diagnostic value in cerebral malaria (CM). To improve our understanding of MR as a predictor of cerebral parasite sequestration in CM we reviewed data from the Blantyre autopsy study.

**Methods:** We performed a retrospective analysis of data collected from a consecutive series of patients presenting to the Pediatric Research Ward at Queen Elizabeth Central Hospital in Blantyre, Malawi between 1996 and 2010. We determined the diagnostic accuracy of MR as a predictor of cerebral parasite sequestration in a cohort of children with fatal CM.

**Results:** Of 84 children included in the study, 65 met the WHO criteria for CM during life. Eighteen (28%) of 65 did not have evidence of cerebral parasite sequestration at autopsy and 17 had an alternative cause of death. MR had a sensitivity of 89% and specificity of 73% to predict sequestration. In a subset of patients with graded retinal assessments, this was improved to 94% and 88% by reclassifying patients with 1-5 hemorrhages in only one eye as retinopathy negative.

**Conclusions:** MR remains the most specific point-of-care test for CM in endemic areas. Its specificity can be improved, without sacrificing sensitivity, by reclassifying patients with less than 5 hemorrhages in one eye only as MR negative.

## Background

The burden of fatal cerebral malaria continues to be high despite decades of high-quality research. In 2022, malaria claimed over 600,000 lives, most being African children aged 0-5 years.^1^ CM is a severe neurological manifestation of the disease characterized by coma and peripheral parasitemia. CM has high mortality even with treatment, and survivors very commonly suffer from neurological injury.^2^

Malarial retinopathy (MR) has diagnostic and prognostic significance in CM.^3,4^ In a landmark autopsy study, Taylor et al. established retinopathy as the only clinical feature which differentiated histopathologically-proven CM from coma of other causes.^3^ In this autopsy cohort (from 1996 – 2000) 23% of cases defined as CM by WHO criteria had no or minimal sequestration and other causes of death, suggesting misclassification. When MR, detected by indirect ophthalmoscopy, was added to the definition of CM, a the sensitivity and specificity for predicting autopsy-established cerebral parasite sequestration improved to 95% and 90%, respectively.^5^ In CM cohorts which include survivors, the proportion of those with non-malarial coma is estimated to be lower at 10%.^6^ There remains a significant proportion of MR negative CM patients for whom parasitemia is a major contributor to coma, perhaps in combination with other factors,.^7^ including co-infection and pre-existing neurological disease.^8,9^

An additional 61 autopsies were performed following the initial analyses in 2004. Here, we provide complete results from the largest autopsy study conducted on pediatric CM cases. We consider these results alongside work published in the two decades following the publication of the findings from the initial autopsy cohort.

## Methods

### Study design

We conducted a retrospective analysis of autopsy cases from the Blantyre Autopsy Study, a case-control study conducted in Queen Elizabeth Central Hospital, Blantyre, Malawi, between 1996 and 2010. We extracted information on clinical diagnosis, histopathological diagnosis, retinopathy status and cause of death. We then established estimates for the performance of MR as a predictor of histopathologically-proven CM.

### Participants

Our cohort was drawn from a consecutive series of patients in coma admitted to the Pediatric Research Ward. In the event of a death, and when the parents or guardians consented, individuals were included. Clinical diagnoses were determined prior to each autopsy according to the following criteria:

#### Cerebral malaria

- Admitting Blantyre Coma Score ≤2, remaining ≤2 following correction of hypoglycaemia, for 30 minutes after the cessation of seizure activity, or 2 hours from admission, and
- *Plasmodium falciparum* parasitemia, and
- No evidence of meningitis following lumbar puncture (i.e., <10 white blood cells/μl, no pathogens seen or cultured)

#### Non-malarial coma

- Admitting Blantyre Coma Score ≤ 2, remaining ≤2 following correction of hypoglycaemia, for 30 minutes after the cessation of seizure activity, or 2 hours from admission, and
- No evidence of malaria infection on a maximum of 4 blood films collected every 6 hours, or
- Non-malarial aetiology of coma identified during life, irrespective of peripheral parasitemia

Detailed descriptions of the patients have been published elsewhere.^3^

### Test methods

#### Index test – presence of malarial retinopathy

Patients underwent retinal examinations on admission by direct and indirect ophthalmoscopy after both eyes were dilated with 1% tropicamide and 2.5% phenylephrine eye drops. Where dilation was insufficient, the drops were repeated. All examinations were performed by either an ophthalmologist or a clinician trained in retinal examination. Ophthalmologists graded MR findings according to a validated grading system and recorded them on a standardised form.^10^ Trained clinicians recorded retinopathy status as either present or absent. Test positivity for clinicians was defined as the presence of any retinal hemorrhages, retinal whitening, or retinal vessel change in either eye. When considering graded ophthalmologist assessments, test positivity was defined as presence of any retinal hemorrhages, retinal whitening (at the macula, fovea, or in any quadrant of the periphery), or retinal vessel change (in any quadrant of the periphery) in either eye. Test negativity was defined as negative for retinal hemorrhages, macular and foveal whitening, and negative peripheral whitening and vessel change with a minimum of three out of four quadrants visualised in both eyes. A total of four ophthalmologist assessments were deemed to be ungradable based upon these criteria. Clinicians performing fundoscopy had access to clinical information about the patient. All fundoscopies were performed during life.

### Reference standard – histopathological evidence of cerebral parasite sequestration

The autopsy procedure has been described in detail elsewhere.^3^ Briefly, in the event of death consent was sought for autopsy by a member of the clinical team, and if granted, performed as soon as possible in the hospital mortuary.

Tissue blocks from standard regions of the cerebrum, brainstem, and cerebellum were fixed in 10% neutral buffered formalin, processed and embedded in paraffin. Haematoxylin and eosin were used to stain 3-5 micron sections. The contents of at least of one hundred perpendicularly cross-sectioned capillaries per section were counted by a single observer. Any blood vessel, circular or oval in profile, with a maximum:minimum diameter of < 2:1, and with at most one visible endothelial cell nucleus in the wall, was considered to be a capillary.

Test positivity was defined as presence of sequestered parasitised red blood cells in the cerebral microvasculature, as determined by a histopathologist. This corresponded to a cut-off value of 23.3% parasitised capillaries, as determined by classification and regression trees.^3^ The pathologist performing histopathological assessment was masked to clinical information and retinopathy findings.

### Statistics

All analyses were conducted in RStudio using R 4.4.0. MR was treated as a binary diagnostic test to predict cerebral parasite sequestration. Sensitivity, specificity, positive predictive value and negative predictive value were calculated using the *testCompareR* package.^11^ Tests were either considered positive, negative or missing. Missing data were handled by complete case analysis. Sample size was dictated by the retrospective nature of this study.

### Ethics

Ethical approval was granted by the research ethics committees at Michigan State University, the University of Liverpool, and the University of Malawi College of Medicine and research was conducted in accordance with the tenets of the Declaration of Helsinki. Written informed consent for participation was obtained from parents or guardians of all particpants.

## Results

Sixty-one autopsies were performed following the initial analysis in 2004, making a total of 103 autopsy examinations (Figure 1). There were 84 cases with retinal data. Sixty-five of these met the WHO criteria for CM during life. Patient demographics are described in Table 1.

**Table 1.** Demographics summarised by presence of cerebral parasite sequestration and presence of malarial retinopathy. * indicates number of participants meeting specified condition (percentage). All other values indicate the median (range). BCS – Blantyre Coma Score; BP – blood pressure; bpm – beats per minute; CSF – cerebrospinal fluid; rpm – respirations per minute.

| Demographics by cerebral sequestration status and malarial retinopathy status |  |  |  |  |
| --- | --- | --- | --- | --- |
| Cerebral parasite sequestration | - | - | + | + |
| Malarial retinopathy | - | + | - | + |
|  | n = 27 | n = 10 | n = 5 | n = 42 |
| Age (months) | 29 (7 - 108) | 20 (6 - 96) | 57.5 (14 - 103) | 30 (6 - 127) |
| No. male (%)* | 16 (59.3) | 6 (60) | 3 (60) | 21 (50) |
| No. fever (%)* | 18 (69.2) | 8 (80) | 4 (80) | 41 (97.6) |
| Fever duration (h) | 48 (2 - 504) | 36 (4 - 72) | 72 (24 - 216) | 48 (4 - 120) |
| Temperature (°C) | 37.5 (32.2 - 42.1) | 37.6 (35 - 40.1) | 37.8 (33.9 - 40.3) | 38.8 (36.5 - 41.9) |
| Heart rate (bpm) | 144 (92 - 180) | 152 (52 - 180) | 144 (120 - 200) | 160 (60 - 233) |
| Respiratory rate (rpm) | 44 (32 - 68) | 42 (20 - 66) | 48 (28 - 50) | 48 (28 - 84) |
| Systolic BP (mmHg) | 100 (74 - 160) | 110 (90 - 175) | 110 (94 - 110) | 102 (70 - 140) |
| Weight (kg) | 10.5 (4 - 26) | 10 (6.5 - 20.5) | 19 (9.1 - 31) | 10 (6.8 - 27) |
| No. BCS 0 (%)* | 14 (51.9) | 3 (30) | 2 (40) | 11 (26.2) |
| No. BCS 1 (%)* | 6 (22.2) | 5 (50) | 2 (40) | 20 (47.6) |
| No. BCS 2 (%)* | 4 (14.8) | 1 (10) | 1 (20) | 11 (26.2) |
| No. BCS > 2 (%)* | 3 (11.1) | 1 (10) | 0 (0) | 0 (0) |
| CSF opening pressure (mmH<sub>2</sub>O) | 215 (20 - 320) | 205.5 (55 - 320) | 170 (80 - 230) | 190 (9 - 320) |
| Time to death (h) | 14 (0 - 168) | 4 (1 - 64) | 18 (5 - 56) | 12.5 (1 - 46) |

**Figure 1.**
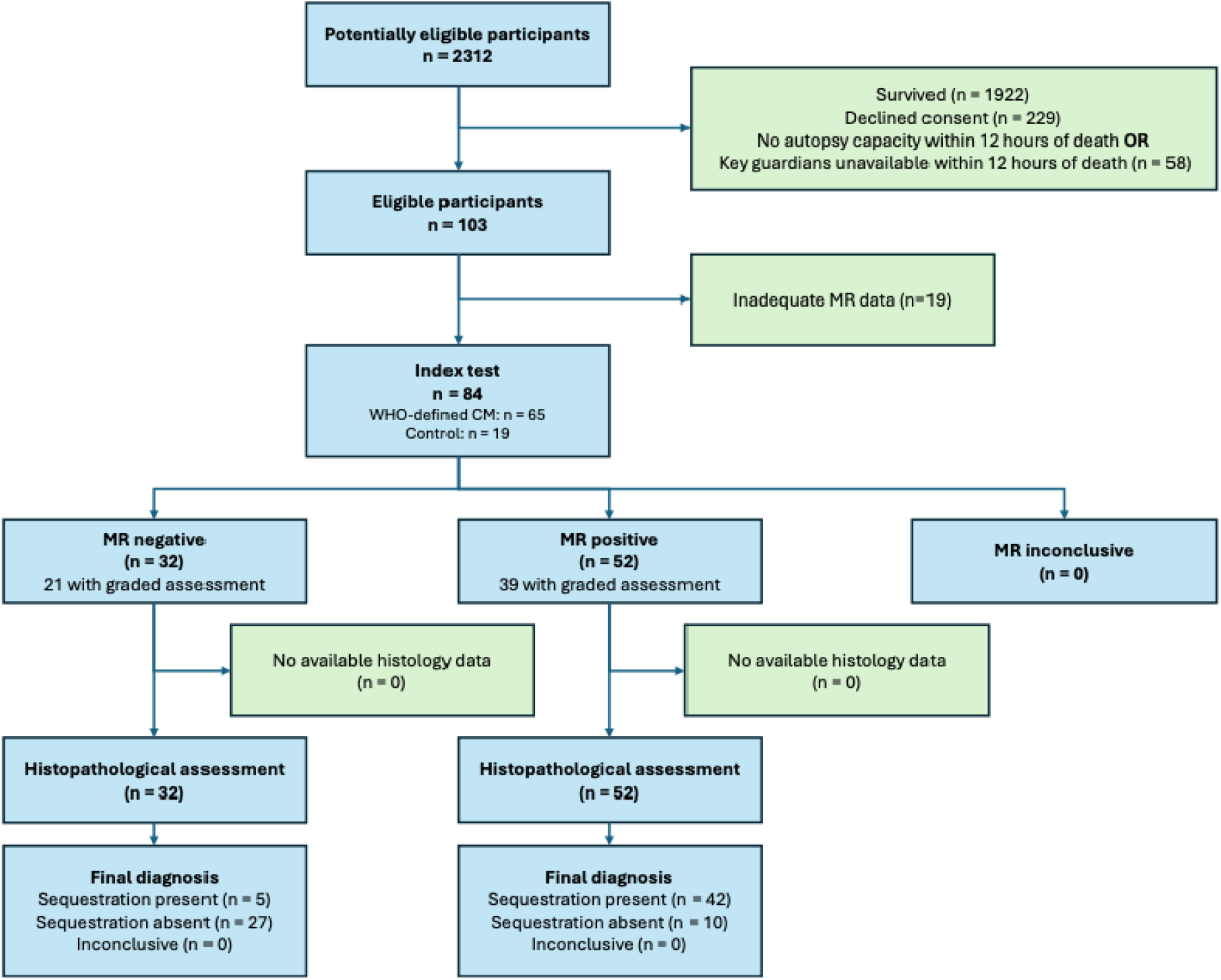
STARD flow chart to describe flow of data through the study. CM – cerebral malaria; MR – malarial retinopathy; WHO – World Health Organization.

We found a sensitivity of 89.4%, which is similar to the initial analysis.^5^ However, the specificity was 73.0%, considerably lower than the initial analysis.^5^ Because changes in specificity are due to false positives, we reviewed the case notes for all false positives to identify the nature of their retinopathy (Supplementary Methods). In four of ten false positives, the only evidence of MR was 1-5 hemorrhages in a single eye. The retinal phenotype for all ten of these patients is described in Supplementary Methods. To test the hypothesis that 1-5 hemorrhages in a single eye is insufficient to predict cerebral parasite sequestration, we reanalysed the 60 patients in whom a graded retinal examination was performed by an ophthalmologist according to the validated system.^8^ Where the only evidence of MR was 1-5 hemorrhages in a single eye, the data were re-coded as no longer constituting a diagnosis of MR. Contingency tables for all analyses are shown in Table 2. The test metrics were compared for the original and the re-coded data (Table 3).

**Table 2.**
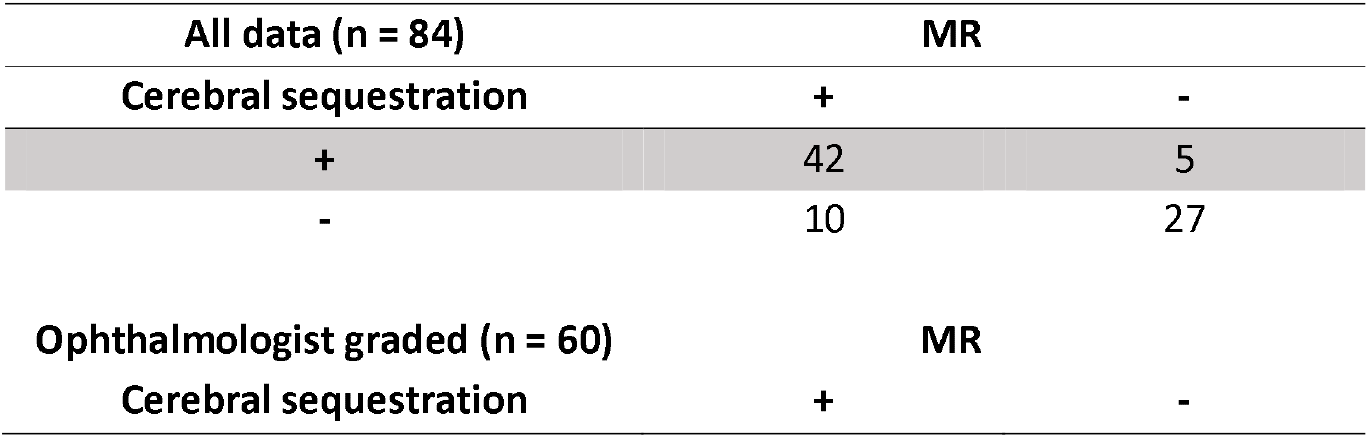
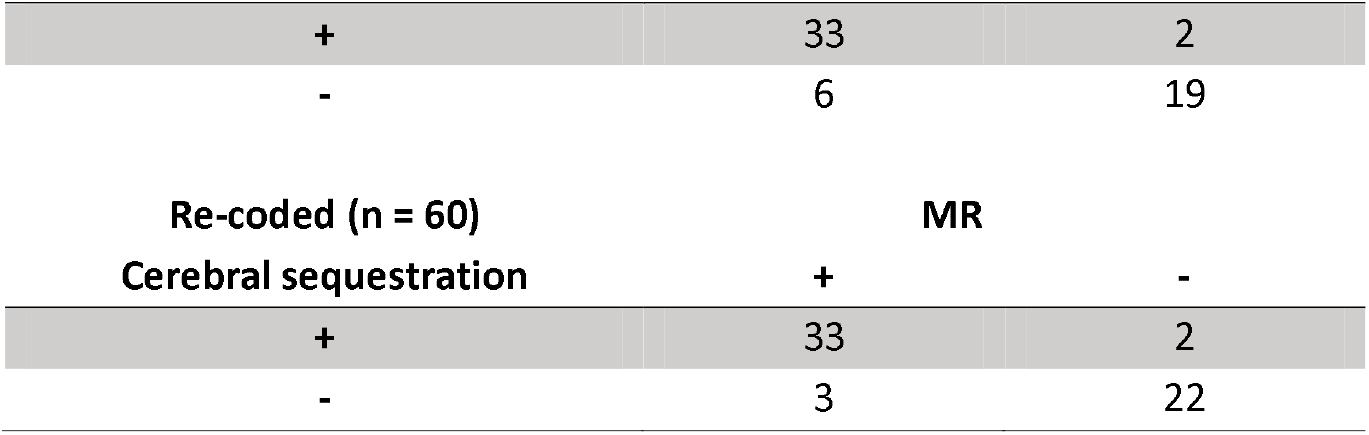
Contingency tables for each of the analyses (whole dataset; ophthalmologist-graded; re-coding 1-5 hemorrhages in a single-eye as MR negative) compared against the gold standard, cerebral parasite sequestration.

**Table 3.** Performance metrics for malarial retinopathy as a predictor of cerebral parasite sequestration. All data includes physician assessment of presence of MR; ophthalmologists performed a complete grading. * p values compare the cohort with graded retinopathy assessments before and after re-coding 1-5 hemorrhages in a single eye as insufficient to define MR. ** Null hypothesis: PLR1 = PLR2 & NLR1 = NLR2, see Supplementary Methods.

| Performance metric | % (Confidence Interval) |  |  | P * |
| --- | --- | --- | --- | --- |
|  | All data (n = 84) | Ophthalmologist graded (n = 60) | Re-coded (n = 60) |  |
| Sensitivity | 89.4 (77.6 – 95.6) | 94.3 (81.7 – 98.7) | 94.3 (81.7 – 98.7) | - |
| Specificity | 73.0 (57.2 – 84.8) | 76.0 (56.8 – 88.8) | 88.0 (70.4 – 96.2) | 0.065 |
| Positive predictive value | 80.8 (71.2 – 87.9) | 84.6 (70.5 – 93.0) | 91.7 (78.5 – 97.4) | 0.34** |
| Negative predictive value | 84.4 (75.2 – 90.7) | 90.5 (71.5 – 97.8) | 91.7 (74.6 – 98.1) | 0.34** |

Re-categorising the participants reduced the number of false positives without a corresponding increase in false negatives, resulting in an improvement in the specificity of MR (p = 0.065). These results indicate that MR can accurately predict cerebral sequestration in comatose children, but also indicate that the specificity of MR can be improved by a definition that excludes cases that have only 1-5 hemorrhages in a single eye.

18 of 65 children (28%) met the clinical case definition for CM but had no evidence of sequestration at autopsy. An alternative cause of death was evident in 17 (Table 4). Four (22%) of these cases had evidence of retinopathy using the original definition, falling to three (17%) using the updated definition.

**Table 4.**
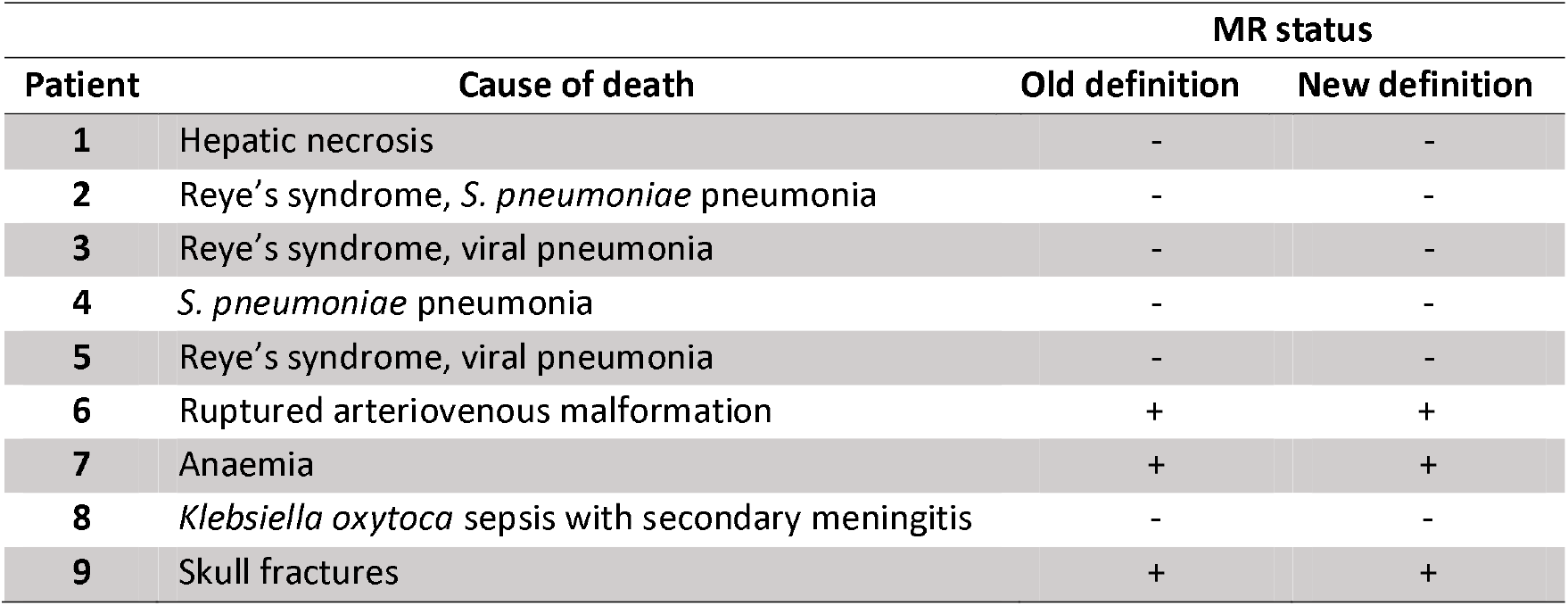
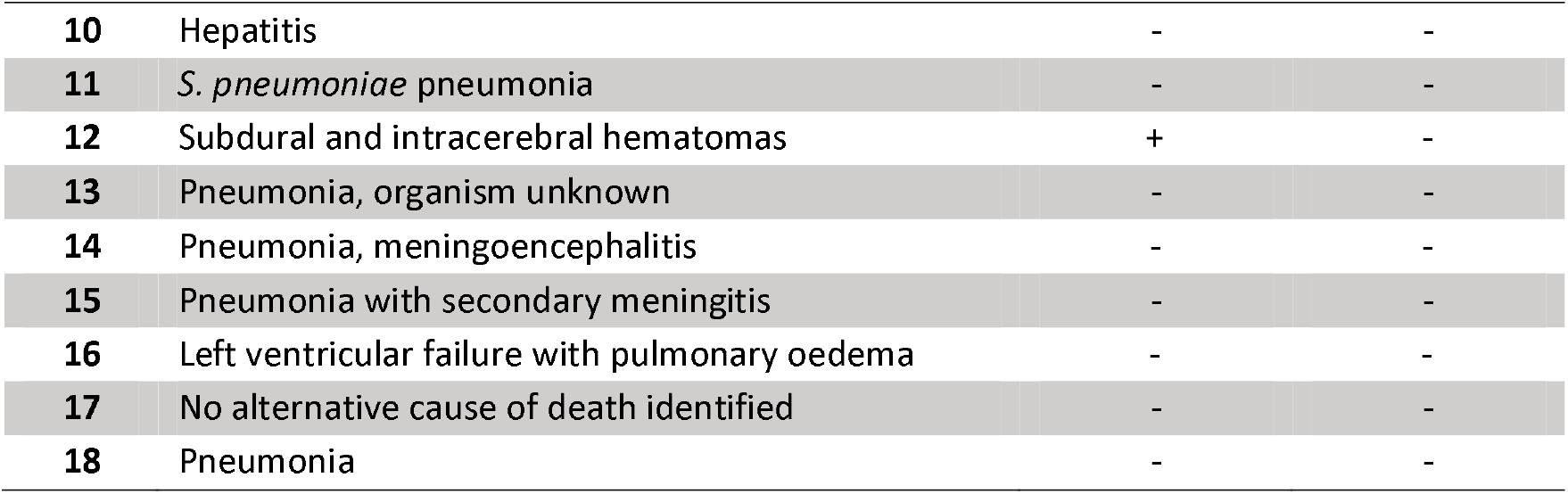
Causes of death and retinopathy status in children meeting the clinical definition of cerebral without cerebral parasite sequestration at autopsy.

| Patient | Cause of death | MR status |  |
| --- | --- | --- | --- |
|  |  | Old definition | New definition |
| 1 | Hepatic necrosis | - | - |
| 2 | Reye's syndrome, *S. pneumoniae* pneumonia | - | - |
| 3 | Reye's syndrome, viral pneumonia | - | - |
| 4 | *S. pneumoniae* pneumonia | - | - |
| 5 | Reye's syndrome, viral pneumonia | - | - |
| 6 | Ruptured arteriovenous malformation | + | + |
| 7 | Anaemia | + | + |
| 8 | *Klebsiella oxytoca* sepsis with secondary meningitis | - | - |
| 9 | Skull fractures | + | + |
| 10 | Hepatitis | - | - |
| 11 | *S. pneumoniae* pneumonia | - | - |
| 12 | Subdural and intracerebral hematomas | + | - |
| 13 | Pneumonia, organism unknown | - | - |
| 14 | Pneumonia, meningoencephalitis | - | - |
| 15 | Pneumonia with secondary meningitis | - | - |
| 16 | Left ventricular failure with pulmonary oedema | - | - |
| 17 | No alternative cause of death identified | - | - |
| 18 | Pneumonia | - | - |

## Discussion

This study confirms that a significant proportion of patients apparently dying of WHO-defined CM have a non-malarial cause of death, and that the absence of MR is an accurate way to identify these patients in life. Reclassifying patients with only 1-5 hemorrhages in one eye improves the specificity of MR as a diagnostic test. These findings further corroborate recent work by our group which showed that retinal hemorrhages alone are insufficient to accurately predict cerebral parasite sequestration in the autopsy cohort, but that retinal whitening and retinal vessel change perform well as predictors.^12^ A major strength of autopsy studies is that they can provide a reference standard by which clinical diagnoses can be validated. However, autopsy studies have inherent selection bias for fatal cases which may compromise the generalizability of their results, and highlight an important question: if surviving MR negative children do not have CM, what do they have?

A large comparison of MR positive and MR negative children found that MR positive children are more likely to present later in their illness, have longer hospital stays, longer coma, and had higher parasite biomass as estimated by parasite protein *Plasmodium falciparum* histidine-rich protein-2 (HRP-2) concentrations.^13^ Rates of bacteraemia did not differ. The authors concluded that CM represents a spectrum of disease, with MR negative children falling at the less severe end. Using more robust statistical methods, the same group subsequently demonstrated increased inflammation in MR positive versus MR negative patients.^14^ The expression of parasite var genes associated with severe malaria is highest in Ugandan children with MR positive CM, followed by those with MR negative CM, severe malarial anemia and asymptomatic parasitemia.^15^ A study by Birbeck et al showed that children with MR negative CM were more likely to have pre-existing developmental problems and family history of epilepsy than controls or MR positive CM cases, suggesting those with MR negative CM have risk factors that make them more susceptible to developing coma.^8^ In conjunction with the autopsy findings, these results support the hypothesis that some cases of MR negative CM are a less severe manifestation of CM or occur in individuals who are more susceptible to coma, while others have a different aetiology altogether.

Modelling studies have attempted to quantify the proportion of MR negative CM children for whom the primary cause of coma is something other than malaria. One recent study found benefit in including retinopathy (alongside platelets and white cell counts) in a model to discriminate true CM.^6^ The researchers employed a Gaussian mixture model using the HRP-2 to classify true CM, based on two assumptions: that HRP-2 concentration accurately predicts cerebral sequestration and that HRP-2 concentration accurately predicts the presence of MR. These assumptions have been validated in the Malawian autopsy cohort and in another cohort of children with CM in Malawi.^16^ HRP-2 concentrations can be quantified in blood, but current point-of-care tests only provide binary results, and these are not useful for differentiating true CM from coma of another cause. The model employed by Yin et al. estimated that 10.7% of comatose, parasitemic children had another cause for their coma.^6^ These results are near to a 7% figure from a natural experiment which leveraged the protection against severe malaria conferred by sickle-cell trait.^7^ Sickle-cell trait was significantly more common in healthy controls than in either MR positive or MR negative children, suggesting that malaria infection contributes to MR negative CM.

Together these data suggest that MR negative CM patients are a heterogenous group, containing patients for whom the major contributing pathogenetic mechanism of their coma is malarial parasitemia. These patients may have a less severe clinical manifestation of CM or have a predisposition to coma. The MR negative CM group also contains children who have a diverse range of non-malarial pathologies (see Table 2). This group ranges from 10% (in children admitted to hospital with WHO-defined CM) to 26% (in fatal cases of WHO-defined CM) justifying comprehensive investigation for other causes of coma in this group and following initiation of treatment for CM. Specifically, those without MR who fail to improve on treatment would benefit from investigations which target the liver, lungs and brain. These findings have important implications for allocation of resources in settings where access may be limited.

There are also important implications for research. Including patients without CM in trials of adjunctive therapies for CM hamper efforts to develop adjunctive treatments. Studies could inadvertently be underpowered to detect a difference between interventional groups if they include non-CM patient. Equally, results in mechanistic studies could be substantially altered by the including patients without CM. Further work is required to definitively characterize MR negative CM, quantifying the proportion of children who have an alternative cause for coma and identifying the factors increasing the susceptibility to coma in some children with malaria infections. These factors may be alteredas malaria vaccine programs are rolled out on larger scales.

This study does have limitations. As previously discussed, autopsy cases carry an inherent selection bias and, though this has been addressed elsewhere in the discussion, the results may not be generalizable to cohorts which include survivors. Additionally, we were unable to compare the original cohort to the more recent cohort directly as, 20 years after the original analysis, some data were irretrievable. Finally, because granular details of the retinal presentation are missing from patients assessed by generalists, the sub-analysis to assess the effect of re-defining the index test as negative when only 1-5 hemorrhages are present in a single eye was necessarily performed using ophthalmologist-graded assessments. In our experience, the interrater reliability between ophthalmologists and trained clinicians in detecting malarial retinopathy is good. Nevertheless, using these data alone it is perhaps more correct to say that we assessed the combined effect of redefining the index test as negative when only 1-5 hemorrhages are present in a single eye and restricting examinations to those performed by ophthalmologists. Going forward, in situations where ophthalmologist examination is impractical, such as large multi-site trials, AI-assisted analysis of images acquired at the bedside may soon be a feasible alternative, provided that the technologies permit good visualisation of the retinal periphery.^17^

At present MR remains the most specific point-of-care test for CM in endemic areas and its specificity can be improved, without sacrificing sensitivity, by reclassifying patients with less than 5 hemorrhages in one eye only as MR negative.

## Supporting information

STARD Checklist

Analysis FIles

## Funding

This work was supported by Wellcome [223502 (KJW) & 222530 (NAVB)].

## Data availability

Data and code required to reproduce this analysis have been provided along with this manuscript (see Supplementary Information).

## Conflicts of interest

The authors declare no conflicts of interest.

## Acknowledgements

We would like to thank the parents and guardians of the children included in this study for their selfless gift, as well as the staff on the Pediatric Research Ward and in the mortuary of Queen Elizabeth Central Hospital, without whom this study would not have been possible.

## Notes

### Competing Interest Statement

The authors have declared no competing interest.

