## Supplementary material for "Re-evaluating malarial retinopathy to improve its diagnostic accuracy in cerebral malaria": Analysis FIles: CompleteAnalysis.pdf

### Re-evaluating retinopathy: data retrieval, validation and statistical analysis

#### 1.0 Introduction

Despite decades of high-quality research, the burden of malaria continues to weigh heavily. In 2021, malaria claimed the lives of over 600000 people, most being African children aged 0-5 years. Cerebral malaria (CM) is a severe neurological manifestation of the disease characterised by coma and peripheral blood parasitaemia. CM mortality is high even with treatment, and survivors very commonly suffer from neurological sequelae.

Malarial retinopathy (MR) has diagnostic and prognostic significance in CM. In a landmark autopsy study, Taylor et al. established retinopathy as the only clinical feature which differentiated histopathologically-proven CM from coma of other causes. Over 20% of this autopsy cohort were misdiagnosed when using the WHO definition of CM alone, with the results suggesting that this could be mitigated by including MR in the definition of CM. A subanalysis of retinal findings established the sensitivity and specificity of MR as 95% and 90%, respectively.

Here we investigate the complete data from the Blantyre autopsy study, collected between 1996 and 2010, to validate and further understand the significance of malarial retinopathy in this cohort.

#### 2.0 Clinical Methods

The Blantyre Autopsy Study was a case-control study conducted in Queen Elizabeth Cental Hospital, Blantyre, Malawi, between 1996 and 2010. Clinical diagnoses were determined prior to each autopsy according to the following criteria:

*Cerebral malaria*

- Admitting Blantyre Coma Score $\leq 2$ and no improvement following correction of hypoglycaemia, within 30 minutes of the cessation of seizure activity, or within 2 hours of admission
- P. falciparum parasitaemia
- No evidence of meningitis following lumbar puncture (i.e., <10 white blood cells/µl, no pathogens seen or cultured)

*Non-malarial coma*

- Admitting Blantyre Coma Score $\leq 2$ and no improvement following correction of hypoglycaemia, within 30 minutes of the cessation of seizure activity, or within 2 hours of admission
- Non-malarial aetiology of coma identified during life, parasitaemic or aparasitaemic (on a maximum of 4 blood films collected every 6 hours), admitting haematocrit $> 15\%$

Detailed descriptions of the patients and autopsies have been published elsewhere (Taylor 2004, Supplmentary Methods).

Ethical approval was granted by the research ethics committees at Michigan State University, the University of Liverpool, and the University of Malawi College of Medicine.

#### 3.0 Analysis Methods

##### 3.1 Cleaning the data

The autopsy data has been archived as an Excel file. This file was loaded into R and some modifications were made in order to facilitate downstream analyses. Analyses are all performed in R Studio using R 4.3.2. For complete `sessionInfo()` see the end of this document.

First we load the necessary packages to complete our analysis and present the results.

```
# install and load packages necessary for analysis and displaying the data
packages <- c("flextable", "readxl", "stringr", "testCompareR", "dplyr", "here")

for (pkg in packages) {
  if (!requireNamespace(pkg, quietly = TRUE)) {
    install.packages(pkg)
  }
  library(pkg, character.only = TRUE)
}
```

Next, we import the data. We can remove all columns except 'Malaria Project Number (MP)', 'CLASS' and 'Ret'. These three are sufficient to define our cases. We rename these columns to make reading the code easier. A sample of the data is displayed.

```r
# import data
tab <- read_xlsx(here("OriginalAutopsyData.xlsx"))

# select and rename essential columns
tab <- tab %>% select(`Malaria Project Number (MP)`, CLASS, Ret)
colnames(tab) <- c("mp_number", "class", "orig_ret")

# view the first 12 rows of the reduced table
head(tab, 12)
```

```
# A tibble: 12 x 3
   mp_number class orig_ret
   <chr>     <chr> <chr>
 1 0550      1     NA
 2 9999      10    NA
 3 9999      10    NA
 4 595       10    NA
 5 628       2     ret
 6 664       2     NA
 7 685       6     no ret
 8 717       7     no ret
 9 742       2     ret
10 999       9     NA
11 770       2     ret
12 787       7     no ret
```

A key for the *class* variable is provided below.

1 = Clinical CM, seq only (> 25% VPC, < 50 pg/100v)
2 = Clinical CM, seq + pigment (> 25% VPC, > 50 pg/100v)
3 = Clinical CM, no seq, no supporting pathology
6 = Clinical severe malaria anemia (no coma, PCV <15%)
7 = Non-malarial encephalopathy, infectious
8 = Non-malarial encephalopathy, non-infectious
9 = Non-malarial cause of death, incidental parasitemia
10 = Indeterminant
11 = Other

In the above table we can see that individuals in whom the MP number is unknown all received a number of 9999. Further examination of the data revealed that one individual received the number 999 likely because of a keystroke error. Having individuals with non-unique identification numbers might disrupt downstream analyses so next, we have to give each of those a unique number. There was only one instance of 999 (out of sequence) so we felt it was reasonable to recode it along with the other non-unique IDs.

```
# replace rows with non-unique PID
replace_rows <- tab$mp_number %in% c(999, 9999)
unique_values <- seq(9999, length.out = sum(replace_rows), by = -1)

tab$mp_number[replace_rows] <- unique_values

# convert to MP#### format
tab <- tab %>%
  mutate(
    mp_number = paste0("MP", str_pad(mp_number, width = 4, pad = 0))
  )

# view the first 12 rows of the reduced table
head(tab, 12)
```

```
# A tibble: 12 x 3
   mp_number class orig_ret
   <chr>     <chr> <chr>
 1 MP0550    1     NA
 2 MP9999    10    NA
 3 MP9998    10    NA
 4 MP0595    10    NA
 5 MP0628    2     ret
 6 MP0664    2     NA
 7 MP0685    6     no ret
 8 MP0717    7     no ret
 9 MP0742    2     ret
10 MP9997    9     NA
11 MP0770    2     ret
12 MP0787    7     no ret
```

Next, we identified several cases where a definitive answer to retinopathy was not provided. These cases were labelled ‘trace whitening’, ‘atypical’ or some variation thereof and ‘1 haemorrhage’. For the current study, these would all be considered as retinopathy positive, so they were recoded as such. It should be noted at this point that none of these cases met the clinical definition of cerebral malaria during life. Therefore, changing all of these to positive will generate false positives. Classifying them as retinopathy negative would generate true negatives only. `NA` values should remain unchanged.

```
tab <- tab %>%
  mutate(
    # remove leading and trailing whitespace from orig_ret column
    orig_ret = str_trim(orig_ret),

    # change all non-specific values to 'ret'
    orig_ret = if_else(
      # check if orig_ret is not 'ret', 'no ret', or NA
      !str_detect(orig_ret, "^ret$|^no ret$|^NA$") & !is.na(orig_ret),
      "ret",   # if condition is TRUE, replace with 'ret'
      orig_ret # if condition is FALSE, keep the original value
    )
  )
```

Next, we can recode 'ret' and 'no ret' to be numerical values 1 and 0, respectively. We can then recode the whole data frame as numeric to faciliate downstream analysis. The `class` value for MP1719 is '7or8' which will introduce an `NA`. As this distinction is not important for these analyses, we changed it to 7.

```
tab <- tab %>%
  mutate(
    # change ret to 1 and no ret to 0
    orig_ret = case_when(
      orig_ret == "ret" ~ 1,
      orig_ret == "no ret" ~ 0
    ),

    # change class for 1719 to 7
    class = if_else(mp_number == "MP1719", "7", as.character(class)),

    # convert data frame to numeric
    orig_ret = as.numeric(orig_ret),
    class = as.numeric(class)
  )
```

Now we have a neat data frame where missing values are coded as `NA` and all other values are numerical, excepting the ID numbers, which are character strings.

True cerebral malaria in this study was defined as histological evidence of sequestration in the cerebral microvasculature. This comprises class 1 and class 2. For more information on what constitutes histological evidence of cerebral parasite sequestration, see the original Nature Medicine paper.

We can define a binary variable which we can later use to establish test metrics using this definition.

```
# define a binary true_cm variable
tab <- tab %>%
  mutate(
    true_cm = if_else(class == 1 | class == 2, # if class 1 or 2
                      1,                        # set value true_cm to 1
                      if_else(class == 10,      # if class 10
                              NA_real_,         # set true_cm to NA
                              0)                # otherwise, set true_cm to 0
                      )
  )
```

The data is now clean. However, the data has been preserved over a long time and may have inadvertently been modified, or there may be discrepancies between the records stored on paper and online. Next, we conducted a chart review and cross-referenced against the original retinopathy findings and online records.

##### 3.2 Retrospective chart review

We conducted a retrospective chart review of all 103 patients included in the autopsy study between 1996 and 2010, extracting information on clinical diagnosis, histopathological diagnosis, retinopathy status. Cause of death was documented at the time of the autopsies and is available in `OriginalAutopsyData.xlsx`. An additional 61 autopsies were performed following the original analyses in 2004.

The data was collected by two reviewers (KJW and AL) and stored in an Excel spreadsheet. The data was imported and the retinopathy data is appended to the data frame.

```
# import data from chart review
chart <- read_xlsx(here("ChartReviewData.xlsx"))

# add column to tab of new chart retinopathy data
tab <- tab %>%
  mutate(
    chart_ret = as.numeric(chart$ret)
  )
```

##### 3.3 Ensuring data quality

###### Checking for differences between the data collected in the chart review and the original data

We checked the records findings of the chart review against the data as it was preserved. A list of case numbers were generated for future review of the paper charts for each mismatched case according to the following criteria:

1) Examination by an ophthalmologist (a graded assessment) is the gold standard and takes precedent over any other findings
2) Where multiple exams have been performed on consecutive days, the first examination should be taken as the truth, except for cases in which the first criterion has been met
3) If the only evidence of retinopathy is circling of the haemorrhage box on the patients admission proforma then

- presence of haemorrhages *is* evidence of malarial retinopathy
- absence of haemorrhages *does not* rule out malarial retinopathy and the entry should be marked `NA`

```
# create a column for disagreements
tab <- tab %>%
  mutate(
    disagree = if_else(
      (is.na(orig_ret) & !is.na(chart_ret)) |
      (!is.na(orig_ret) & is.na(chart_ret)) |
      (orig_ret != chart_ret), 1, 0)
  )

disagree <- tab %>%
  filter(disagree == 1) %>%
  pull(mp_number)

# print the results
cat(
"The following cases disagree:", disagree,
"\nThere are", disagree %>% length(), "disagreements.",
sep = " "
)
```

```
The following cases disagree: MP0550 MP0628 MP0664 MP0787 MP0795 MP9996 MP1354 MP1528 MP1551
There are 15 disagreements.
```

##### Evaluating the online records

Next, we established if there were any ophthalmology exams recorded in an online database of retinopathy findings, which is a digitised version of the paper standardised grading forms routinely used to record this data. Digitisation included removing, scanning and inputting the data, then returning the forms to the notes. It is possible that some forms went missing during this process, making the database the only enduring record of those examinations. As we are prioritising detailed ophthalmic assessment, these forms are at the top of our data quality hierarchy.

We made a decision to include any graded assessment where more than 50% of the retina was seen.

*NB. The data provided with this file includes only the retinopathy data required to reproduce the analysis. Access to the full retinopathy database should be requested from the Blantyre Malaria Project who maintain the archive. Contact Kyle Wilson or Karl Seydel for further information.*

```
# import graded retinopathy database
rtd <- read.csv(here("RetinopathyData.csv"))

# BELOW COMMENTED OUT - ONLY USE WITH COMPLETE DATASET
# make cases consistent and remove whitespace
# rtd$mpid <- toupper(rtd$mpid)
# rtd$mpid <- gsub(" ", "", rtd$mpid)

# filter cases in the autopsy study
# rtd <- rtd %>% filter(rtd$mpid %in% tab$mp_number)

# clean to include only specific columns of interest
rtd <- rtd %>% select(1:3,6:7,12:15,24:27,92:95,96,98)
colnames(rtd) <- c("mp_number", "haem_r", "haem_l", "pap_r", "pap_l",
                   "mac_wh_r", "mac_wh_l", "fov_wh_r", "fov_wh_l",
                   "per_wh_r", "per_wh_l", "wh_qu_seen_r", "wh_qu_seen_l",
                   "quad_vc_r", "quad_vc_l", "vc_qu_seen_r", "vc_qu_seen_l",
                   "score", "doc")

# save full_ret data frame for later analysis
full_ret <- rtd

# use redcap codebook to recode to 1 (present), 0 (absent) or NA for each sign
# allow cases where greater than 50% retina seen in both eyes
rtd <- rtd %>% transmute(
  mp_number = mp_number,
```

```r
  haem = if_else(haem_r > 0 | haem_l > 0, 1,
                 if_else(haem_r == 0 & haem_l == 0, 0, NA_real_)),
  white = if_else(mac_wh_r > 0 | mac_wh_l > 0 | fov_wh_r > 0 | fov_wh_l > 0 |
                  per_wh_r > 0 | per_wh_l > 0, 1,
                  if_else(mac_wh_r == 0 & mac_wh_l == 0 & fov_wh_r == 0 &
                          fov_wh_l == 0 & per_wh_r == 0 & per_wh_l == 0 &
                          wh_qu_seen_r >= 3 & wh_qu_seen_l >= 3, 0, NA_real_)),
  vc = if_else(quad_vc_r %in% 1:4 | quad_vc_l %in% 1:4, 1,
               if_else(quad_vc_r == 0 & quad_vc_l == 0 &
                       (vc_qu_seen_r == 3 | vc_qu_seen_r == 4) &
                       (vc_qu_seen_l == 3 | vc_qu_seen_l == 4), 0, NA_real_))
  )

# add a column for overall retinopathy status
rtd$ophth_ret[rtd$haem > 0 | rtd$white > 0 | rtd$vc > 0] <- 1
rtd$ophth_ret[rtd$haem == 0 & rtd$white == 0 & rtd$vc == 0] <- 0
```

Now, we need to join the retinopathy status from the chart review so we can look for mismatches.

```r
# join dataframe column
rtd <- rtd %>%
  left_join(tab %>% select(mp_number, chart_ret),
            by = join_by(mp_number == mp_number))

# find differences
rtd$diff <- case_when(
  rtd$ophth_ret != rtd$chart_ret ~ 1,
  is.na(rtd$chart_ret) & !(is.na(rtd$ophth_ret)) ~ 1,
  !(is.na(rtd$chart_ret)) & is.na(rtd$ophth_ret) ~ 1,
  TRUE ~ 0
)

print(rtd %>% filter(diff == 1) %>% select(mp_number, chart_ret, ophth_ret))
```

```
  mp_number chart_ret ophth_ret
1    MP0871         0        NA
2    MP1223        NA         1
3    MP1388         0         1
4    MP1777         0        NA
5    MP2011        NA         0
```

```
6    MP2031         0        NA
7    MP2083        NA         0
```

None of these cases appear in our list of disagreements, which we can confirm computationally.

We should therefore update each of these cases so that the **true** value reflects the graded assessment, as per our hierarchy of data.

```r
# confirm graded vs. chart do not appear in chart vs. orig disagreements
TRUE %in% (disagree %in% rtd$diffs)
```

```
[1] FALSE
```

```r
# returns FALSE if no matches between disagree and diffs

# update the values as above
tab <- tab %>%
  mutate(final_ret = case_when(
    mp_number %in% c("MP0871", "MP1777", "MP2031") ~ NA_real_,
    mp_number %in% c("MP1223", "MP1388") ~ 1,
    mp_number %in% c("MP2011", "MP2083") ~ 0,
    TRUE ~ chart_ret  # keep original values for other cases
  ))
```

Next, we remove the cases where there is no disagreement between the **chart review** and the **graded assessments** from our list of disagreements which need to be reviewed, as the grading verifies the findings of the chart review. As no discrepancies between the chart review and the graded assessments appear in our list of disagreements from earlier, we can just use a simple function to compare the vectors.

```r
# create a vector of differences remaining
rem_disagree <- setdiff(disagree, rtd$mp_number)

cat(
"The following cases disagree:", rem_disagree,
"\nThere are", rem_disagree %>% length(), "disagreements.",
sep = " "
)
```

```
The following cases disagree: MP0550 MP0628 MP0664 MP0787 MP0795 MP9996 MP1528 MP1551 MP1682
There are 13 disagreements.
```

Each of these cases were reviewed again by two authors (KJW and KBS). The reasons for each difference were documented, along with whether the chart review values needed to be changed. This has been summarised in Table 1.

```
# nb. some steps here may seem unnecessary, but they ensure nothing was missed
# during steps taken in Excel

# create a data frame with 3 columns
table1 <- as.data.frame(rem_disagree)

# import comments and change indicators
comments <- read_xlsx(here("ChartReviewData.xlsx"), sheet = 2)
comments <- comments %>%
  mutate(
    mp_number = paste0("MP", str_pad(mp_number, 4, pad = 0))
  )

# join to table1
table1 <- left_join(table1, comments %>% select(mp_number, short_comments),
                    by = join_by(rem_disagree == mp_number))

# add change column
changed <- "MP2131"

table1 <- table1 %>%
  mutate(change = if_else(rem_disagree %in% changed, "Yes", "No"))

# format for printing
colnames(table1) <- c("MP number", "Comments", "Changed")

# print as a flextable
ft1 <- flextable(table1)
ft1 %>% # maybe print in appendix
  autofit() %>%
  align(align = "center", part = "header") %>%
  align(align = "center", part = "body", j = c(1,3))
```

| MP number | Comments | Changed |
|---|---|---|
| MP0550 | Positive - documented on cover sheet (haemorrhage) | No |
| MP0628 | Negative - documented in notes | No |

| MP number | Comments | Changed |
|---|---|---|
| MP0664 | Positive - documented on cover sheet (haemorrhage) | No |
| MP0787 | Positive - documented in notes | No |
| MP0795 | Negative - consensus decision - notes unclear | No |
| MP9996 | Unable to retrieve case notes | No |
| MP1528 | NA - consensus decision - notes unclear | No |
| MP1551 | Positive - documented on cover sheet (haemorrhage) | No |
| MP1682 | Positive - documented on cover sheet (haemorrhage) | No |
| MP1971 | Negative - documented in notes | No |
| MP1979 | Negative - documented in notes | No |
| MP2131 | NA - consensus decision - limited exam | Yes |
| MP2138 | Positive - documented in notes | No |

To complete the final retinopathy classification, we should update the value from the table which is marked for change.

```
# update the values as above
tab <- tab %>%
  mutate(final_ret = case_when(
    mp_number == "MP2131" ~ NA_real_,
    TRUE ~ final_ret  # keep original values for other cases
  ))
```

Finally, we can create a cleaner data frame for downstream analysis which contains all of the patients with the MP number, final retinopathy status and true cerebral malaria status. We can also create a data

```
# select necessary columns
dat <- tab %>% select(mp_number, final_ret, true_cm)

# clear the work space
rm(list = setdiff(ls(), c("dat", "full_ret")))
```

##### 3.4 Statistical analyses

Next we need to examine the test metrics for the retinopathy findings as determined by the original reviewers and the chart review that we conducted.

Analyses were conducted using the R package `testCompareR`. By default, this includes correction for multiple comparisons. Accordingly, reported p values have been adjusted using the Holm method.

```
# calculate test metrics with testCompareR package, print with interpretR()
all <- dat %>%
  filter(complete.cases(final_ret) & complete.cases(true_cm))

res_all <- summariseR(data.frame(all$final_ret, all$true_cm)) %>%
  interpretR()
```

```
--------------------------------------------------------------------------------
CONTINGENCY TABLES
--------------------------------------------------------------------------------

           Test
True Status Positive Negative
   Positive       42        5
   Negative       10       27

--------------------------------------------------------------------------------
PREVALENCE (%)
--------------------------------------------------------------------------------

           Estimate  SE Lower CI Upper CI
Prevalence       56 5.4     45.3     66.1

--------------------------------------------------------------------------------
DIAGNOSTIC ACCURACIES
--------------------------------------------------------------------------------

            Estimate  SE Lower CI Upper CI
Sensitivity     89.4 4.5     77.6     95.6
Specificity     73.0 7.3     57.2     84.8

--------------------------------------------------------------------------------
```

```
PREDICTIVE VALUES
---------------------------------------------------------------------------------

    Estimate  SE Lower CI Upper CI
PPV     80.8 0.6     71.2     87.9
NPV     84.4 0.7     75.2     90.7

---------------------------------------------------------------------------------
LIKELIHOOD RATIOS
---------------------------------------------------------------------------------

    Estimate  SE Lower CI Upper CI
PLR      3.3 0.9      2.0      5.7
NLR      0.1 0.1      0.1      0.3

---------------------------------------------------------------------------------
COHEN'S KAPPA
---------------------------------------------------------------------------------

0.632
```

##### Re-defining retinopathy such that 1-5 haemorrhages in one eye is not considered predictive of cerebral parasite sequestration

Lower specificity than originally expected prompted us to take a closer look at all of the cases that were false positives, that is to say they had no evidence of cerebral sequestration of malarial parasites, but they did have evidence of malarial retinopathy. These case were identified from all complete cases.

```
all %>% filter(true_cm == 0 & final_ret == 1) %>% pull(mp_number)
```

```
 [1] "MP0787" "MP0899" "MP1182" "MP1388" "MP1501" "MP1551" "MP1665" "MP1682"
 [9] "MP2021" "MP2230"
```

Records for these cases were examined to try and get a more complete understanding of why they were false positives.

For the cases where retinopathy was identified, but the patient had no evidence of sequestration we discovered:

- MP0787 had a single haemorrhage and macular oedema in 1 eye, diagnosis
- MP0899 had a single spot of retinal whitening in 1 eye, diagnosis

- MP1182 had 1-5 haemorrhages only in 1 eye, diagnosis clinical severe malaria without coma
- MP1388 had peripheral whitening only in both eyes, diagnosis infectious encephalopathy (non-malaria)
- MP1501 had 1-5 haemorrhages only in 1 eye, diagnosis non-infectious encephalopathy
- MP1551 had haemorrhages only, many in right eye, diagnosis clinical CM without sufficient sequestration for diagnosis
- MP1665 had peripheral whitening only in both eyes, diagnosis clinical CM without sufficient sequestration for diagnosis
- MP1682 had MR with all signs, diagnosis CM without sufficient sequestration for diagnosis
- MP2021 had 1-5 haemorrhages only in 1 eye, diagnosis clinical CM without sufficient sequestration for diagnosis
- MP2230 had 1-5 haemorrhages in 1 eye and 6-20 haemorrhages in the other eye, diagnosis non-infectious encephalopathy

In several cases, the only findings were a small number of haemorrhages in one eye only.

To test the hypothesis that redefining retinopathy such that 1-5 haemorrhages in one eye only is insufficient to predict cerebral parasite sequestration we first needed to define a list of patients with graded ophthalmology assessment and calculate the test metrics in that subset. We used the database of ophthalmic grading forms to identify patients that had graded ophthalmology assessment.

In the database codebook haemorrhages for each eye are coded as either 0 (absent), 1 (1-5) or 2,3 or 7 (increasing severity greater than 5 haemorrhages). Negative numbers indicate missing data. The codebook is provided in the associated files.

We coded each eye individually for haemorrhages, as well as a global score for whitening and vessel change.

```
# use redcap codebook to recode to 1 (present), 0 (absent) or NA for each sign
# allow cases where greater than 50% retina seen in both eyes
full_ret <- full_ret %>% transmute(
  mp_number = mp_number,
  haem_r = if_else(haem_r == 1, 1,
               if_else(haem_r >= 2, 2,
               if_else(haem_r == 0, 0, NA_real_))),
  haem_l = if_else(haem_l == 1, 1,
               if_else(haem_l >= 2, 2,
               if_else(haem_l == 0, 0, NA_real_))),
  white = if_else(mac_wh_r > 0 | mac_wh_l > 0 | fov_wh_r > 0 | fov_wh_l > 0 |
                  per_wh_r > 0 | per_wh_l > 0, 1,
                  if_else(mac_wh_r == 0 & mac_wh_l == 0 & fov_wh_r == 0 &
```

```r
                             fov_wh_l == 0 & per_wh_r == 0 & per_wh_l == 0 &
                             wh_qu_seen_r >= 3 & wh_qu_seen_l >= 3, 0, NA_real_)),
  vc = if_else(quad_vc_r %in% 1:4 | quad_vc_l %in% 1:4, 1,
               if_else(quad_vc_r == 0 & quad_vc_l == 0 &
                       (vc_qu_seen_r == 3 | vc_qu_seen_r == 4) &
                       (vc_qu_seen_l == 3 | vc_qu_seen_l == 4), 0, NA_real_))
  )

# define retinopathy as any positive finding in either eye
full_ret$ophth_ret[full_ret$haem_r > 0 | full_ret$haem_l > 0|
              full_ret$white > 0 | full_ret$vc > 0] <- 1
full_ret$ophth_ret[full_ret$haem_r == 0 & full_ret$haem_l == 0 &
              full_ret$white == 0 & full_ret$vc == 0] <- 0

true_cm <- dat %>% filter(mp_number %in% full_ret$mp_number) %>% pull(true_cm)
full_ret <- full_ret %>%
  mutate(
    true_cm = true_cm
  )

# identify cases where there are 1-5 haemorrhages only in one eye
one_to_five <- full_ret %>% filter((haem_r == 1 & haem_l == 0 &
                               white == 0 & vc == 0)|
                              (haem_l == 1 & haem_r == 0 &
                               white == 0 & vc == 0)) %>% pull(mp_number)

# identify cases where there are 1-5 haemorrhages only in both eyes
both <- full_ret %>% filter(haem_r == 1 & haem_l == 1 &
                       white == 0 & vc == 0) %>% pull(mp_number) # no cases
```

Finally, we recoded the retinopathy status for all of those cases where the only evidence of retinopathy was one to five harmorrhages in one eye only to be negative. We then re-run the analysis comparing all parameters using the `testCompareR` package.

```r
full_ret <- full_ret %>%
  mutate(
    recoded = if_else(mp_number %in% one_to_five, # if 1-5 haemorrhages
                      0,                          # recode as no retinopathy
                      ophth_ret)                  # otherwise, no change
  )

# create the data frame for testCompareR
```

```r
ret_recode <- full_ret %>%
  filter(complete.cases(ophth_ret) & complete.cases(true_cm) &
           complete.cases(recoded)) %>%
  select(ophth_ret, recoded, true_cm)

# compare original and recoded data
compareR(ret_recode) %>% interpretR()
```

```
WARNING:
Zeros exist in contingency table. Tests may return NA/NaN.

--------------------------------------------------------------------------------
CONTINGENCY TABLES
--------------------------------------------------------------------------------

True Status - POSITIVE
          Test 2
Test 1     Positive Negative
  Positive       33        0
  Negative        0        2

True Status - NEGATIVE
          Test 2
Test 1     Positive Negative
  Positive        3        3
  Negative        0       19

Gold standard vs. Test 1
             Test 1
Gold standard Positive Negative
     Positive       33        2
     Negative        6       19

Gold standard vs. Test 2
             Test 2
Gold standard Positive Negative
     Positive       33        2
     Negative        3       22

--------------------------------------------------------------------------------
PREVALENCE (%)
```

```
--------------------------------------------------------------------------------

           Estimate  SE Lower CI Upper CI
Prevalence     58.3 6.4     45.8       70

--------------------------------------------------------------------------------
DIAGNOSTIC ACCURACIES
--------------------------------------------------------------------------------

 Test 1 (%)
            Estimate  SE Lower CI Upper CI
Sensitivity     94.3 3.9     81.7     98.7
Specificity     76.0 8.5     56.8     88.8

 Test 2 (%)
            Estimate  SE Lower CI Upper CI
Sensitivity     94.3 3.9     81.7     98.7
Specificity     88.0 6.5     70.4     96.2

Global hypothesis testing was not or could not be performed.
This is usually because:
* prevalence is <10% and total number of participants < 100
* tests have identical performance
* there are many zeros in the contingency table
* Youden Index is < 0

Consider the quality of your data and/or tests. Individual tests have been
attempted below.

Investigating individual differences

Null Hypothesis 1: Se1 = Se2
Test statistic:  NaN  Adjusted p value:  NaN
NAs or NaNs in individual tests.

This is usually because:
* tests have identical performance
* there are many zeros in the contingency table
* Youden Index is < 0

Consider the quality of your data and/or tests.

Null Hypothesis 2: Sp1 = Sp2
```

```
Test statistic:  3.409091  Adjusted p value:  0.06483816

--------------------------------------------------------------------------------
PREDICTIVE VALUES
--------------------------------------------------------------------------------

 Test 1 (%)
    Estimate  SE Lower CI Upper CI
PPV     84.6 5.8     70.5     93.0
NPV     90.5 6.4     71.5     97.8

 Test 2 (%)
    Estimate  SE Lower CI Upper CI
PPV     91.7 4.6     78.5     97.4
NPV     91.7 5.6     74.6     98.1

Global Null Hypothesis: PPV1 = PPV2 & NPV1 = NPV2
Test statistic:  3.257746  Adjusted p value:  0.3393162
Do not reject the global null hypothesis. Difference in predictive values is NOT significant

--------------------------------------------------------------------------------
LIKELIHOOD RATIOS
--------------------------------------------------------------------------------

 Test 1 (%)
    Estimate  SE Lower CI Upper CI
PLR      3.9 1.4      2.1      7.7
NLR      0.1 0.1      0.0      0.2

 Test 2 (%)
    Estimate  SE Lower CI Upper CI
PLR      7.9 4.3      3.1     19.0
NLR      0.1 0.0      0.0      0.2

Global Null Hypothesis: PLR1 = PLR2 & NLR1 = NLR2
Test statistic:  3.54794  Adjusted p value:  0.3393162
Do not reject the global null hypothesis. Difference in likelihood ratios is NOT significant
```

#### 4.0 Conclusions

We present the complete analysis of retinopathy data from the Blantyre autopsy study for the first time. This data provides novel insights into which retinal findings are predictive of cerebral parasite sequestration.

Specifically, 1-5 haemorrhages in a single eye without any other evidence of retinopathy does *not* appear to be predictive of cerebral parasite sequestration in our autopsy cohort. Further details are available in the main paper.

```r
sessionInfo()
```

```
R version 4.4.0 (2024-04-24)
Platform: aarch64-apple-darwin20
Running under: macOS Ventura 13.5.1

Matrix products: default
BLAS:   /Library/Frameworks/R.framework/Versions/4.4-arm64/Resources/lib/libRblas.0.dylib
LAPACK: /Library/Frameworks/R.framework/Versions/4.4-arm64/Resources/lib/libRlapack.dylib;  [illegible]

locale:
[1] en_US.UTF-8/en_US.UTF-8/en_US.UTF-8/C/en_US.UTF-8/en_US.UTF-8

time zone: Europe/London
tzcode source: internal

attached base packages:
[1] stats     graphics  grDevices utils     datasets  methods   base

other attached packages:
[1] here_1.0.1         dplyr_1.1.4        testCompareR_1.0.3 stringr_1.5.1
[5] readxl_1.4.3       flextable_0.9.6

loaded via a namespace (and not attached):
 [1] utf8_1.2.4              generics_0.1.3          fontLiberation_0.1.0
 [4] xml2_1.3.6              stringi_1.8.4           httpcode_0.3.0
 [7] digest_0.6.36           magrittr_2.0.3          evaluate_0.24.0
[10] grid_4.4.0              fastmap_1.2.0           rprojroot_2.0.4
[13] cellranger_1.1.0        jsonlite_1.8.8          zip_2.3.1
[16] crul_1.5.0              promises_1.3.0          fansi_1.0.6
[19] fontBitstreamVera_0.1.1 textshaping_0.4.0       cli_3.6.3
[22] shiny_1.8.1.1           rlang_1.1.4             fontquiver_0.2.1
[25] crayon_1.5.3            withr_3.0.0             gfonts_0.2.0
[28] yaml_2.3.9              gdtools_0.3.7           tools_4.4.0
[31] officer_0.6.6           uuid_1.2-0              httpuv_1.6.15
[34] curl_5.2.1              vctrs_0.6.5             R6_2.5.1
[37] mime_0.12               lifecycle_1.0.4         ragg_1.3.2
```

```
[40] pkgconfig_2.0.3      pillar_1.9.0        later_1.3.2
[43] data.table_1.15.4    glue_1.7.0          Rcpp_1.0.13
[46] systemfonts_1.1.0    tidyselect_1.2.1    tibble_3.2.1
[49] xfun_0.46            rstudioapi_0.16.0   knitr_1.48
[52] xtable_1.8-6         htmltools_0.5.8.1   rmarkdown_2.27
[55] compiler_4.4.0       askpass_1.2.0       openssl_2.2.0
```
